# ICD-10 based syndromic surveillance enables robust estimation of burden of severe COVID-19 requiring hospitalization and intensive care treatment

**DOI:** 10.1101/2022.02.11.22269594

**Authors:** Kristin Tolksdorf, Walter Haas, Ekkehard Schuler, Lothar H. Wieler, Julia Schilling, Osamah Hamouda, Michaela Diercke, Silke Buda

## Abstract

**Objective:** The emergence of coronavirus disease 2019 (COVID-19) required countries to establish COVID-19 surveillance by adapting existing systems, such as mandatory notification and syndromic surveillance systems. We estimated age-specific COVID-19 hospitalization and intensive care unit (ICU) burden from existing severe acute respiratory infections (SARI) surveillance and compared the results to COVID-19 notification data.

**Methods:** Using data on SARI cases with ICD-10 diagnosis codes for COVID-19 (COVID-SARI) from the ICD-10 based SARI sentinel, we estimated age-specific incidences for COVID-SARI hospitalization and ICU for the first five COVID-19 waves in Germany and compared these to incidences from notification data on COVID-19 cases using relative change Δ*r* at the peak of each wave.

**Findings:** The COVID-SARI incidence from sentinel data matched the notified COVID-19 hospitalization incidence in the first wave with Δ*r*=6% but was higher during second to fourth wave (Δ_*r*_ =20% to 39%). In the fifth wave, the COVID-SARI incidence was lower than the notified COVID-19 hospitalization incidence (Δ_*r*_ =-39%). For all waves and all age groups, the ICU incidence estimated from COVID-SARI was more than twice the estimation from notification data.

**Conclusion:** The use of validated SARI sentinel data adds robust and important information for assessing the true disease burden of severe COVID-19. Mandatory notifications of COVID-19 for hospital and ICU admission may underestimate (work overload in local health authorities) or overestimate (hospital admission for other reasons than the laboratory-confirmed SARS-CoV-2 infection) disease burden. Syndromic ICD-10 based SARI surveillance enables sustainable cross-pathogen surveillance for seasonal epidemics and pandemic preparedness of respiratory viral diseases.

## Introduction

After the first cases of coronavirus disease 2019 (COVID-19) had been identified in Europe at the end of January, 2020, reporting systems had to be adapted to the newly emerging pandemic (1). The World Health Organization (WHO) supported countries worldwide with guidance on COVID-19 surveillance, indicating that regular revision would be necessary and country-specific adaptions may be needed (2). In addition to active case finding and mandatory reporting of COVID-19 cases, the continued use of existing syndromic surveillance systems for respiratory diseases was recommended (3). Syndromic surveillance has been used to inform pandemic influenza severity assessments (PISA), providing information on transmissibility, seriousness of disease and impact on health systems (4). These indicators, developed in preparation for a potential new influenza pandemic, can be adapted for the COVID-19 pandemic as well.

Considering the repeated emergence of new concerning variants of the SARS-CoV-2 virus and the waning of vaccine-acquired or natural immunity, the ongoing surveillance of severe COVID-19 cases of all age groups is essential to assess disease seriousness and to identify risk groups (4-6). However, the actual burden of severe COVID-19 cases is difficult to determine, as valid data on hospitalized cases is neither easy to get nor easy to interpret.

The mandatory notification and reporting system in Germany collects PCR-confirmed SARS-CoV-2 infections. The information is sent from the laboratory to the local health authority (LHA) in charge, where all further epidemiological information including information on hospitalization, treatment at intensive care unit (ICU) and death of COVID-19 cases have to be investigated. However, information on hospitalization was not always possible to obtain, since continuous follow-up of cases by the local public health authorities was required. Prior analyses of notification data from the first COVID-19 wave have shown, that 13% of COVID-19 cases lacked information on hospitalization status (7). Among cases with known hospitalization status, about 10% of cases had no information on intensive care and about 50% did not have information on discharge of hospital. In July 2021, a new regulation was implemented to improve completeness of notification data regarding hospitalization associated with a SARS-CoV-2 infection. Hospitals had to notify every hospitalized patient with SARS-CoV-2 infection to the respective LHA. But especially during the peaks of the waves, lacking information on hospitalization and especially on ICU treatment was still expected. Thus, there was urgent need for additional, reliable information on hospitalized patients to validate notification data on severe cases and to give a robust estimation of COVID-19 hospitalization and intensive care burden.

Here, we analyzed data from the ICD-10 based hospital sentinel for severe acute respiratory infections (ICOSARI) combined with ICD-10 codes on COVID-19 with laboratory confirmation to estimate age-specific COVID-19 hospitalization and intensive care incidences in accordance to WHO recommendations (2-4). Using data from five COVID-19 waves in Germany, we provide weekly estimates of severe COVID-19 burden for six age groups to show the robustness of our method and point out the relevance of this important additional data source for a thorough understanding of the actual pandemic situation and expected trends in disease severity.

## Methods

### Data sources

#### ICOSARI sentinel

We use data from the syndromic hospital surveillance system ICOSARI (ICD-10 based hospital sentinel for severe acute respiratory infections), which was established in 2015 in cooperation with a large hospital network (8). The ICOSARI sentinel comprises 71 acute care hospitals in 13 of 16 federal states and covers about 6% of hospitalized patients in Germany. Within the participating hospitals, case-based data on admitted patients are routinely recorded. Data from the hospitals are collected and validated centrally within a data center. From there, anonymized case-based reports on all-cause admissions (***denominator dataset***) as well as on all patients admitted with any respiratory diagnosis including ICD-10 codes for primary and secondary diagnoses (***numerator dataset***) are transferred weekly to the national health institute. Weekly analyses are age-stratified and focused on SARI cases (ICD-10 primary diagnosis codes J09-J22, Basic Case Definition BCD, and ICD-10 primary or secondary diagnosis codes J09-J22, Sensitive Case Definition SCD), which are extracted from the ***numerator dataset*** (9, 10). The system has been validated and was described in detail in 2017 (8). Since January 2017, the ICOSARI system is an integral component of the syndromic surveillance for acute respiratory infections, in combination with syndromic surveillance systems on population and on out-patient level (11, 12).

At the end of February, 2020, an ICD-10 diagnosis code for COVID-19 with laboratory confirmation (ICD-10 secondary diagnosis code U07.1!) was introduced and rapidly implemented in the ICOSARI sentinel (9, 10). Since March, 2020, we have enhanced the SARI monitoring to daily data transfers including preliminary ICD-10 codes of hospitalized patients. In particular, we have expanded our analyses to SARI cases (SCD) with ICD-10 code for laboratory confirmed COVID-19 (***COVID-SARI cases***)(13). We defined a severe COVID-19 case (hospitalized due to the SARS-CoV-2 infection) as a COVID-SARI case.

From the ICOSARI sentinel, we used the yearly number of patient admissions and the variables age, sex and 2-digit postal code from the ***denominator dataset*** to estimate the catchment population. For the estimation of COVID-SARI incidence, we additionally used the variables age, ICU admission and ICD-10 codes from the ***numerator dataset***. We included SARI cases (primary diagnosis codes J09 to J22) from week 1/2015 to week 52/2019 to validate the method. We included COVID-SARI cases (primary or secondary diagnosis codes J09 to J22 with secondary diagnosis code U07.1! for laboratory confirmed COVID-19) from week 10/2020 to week 20/2022 to estimate weekly incidences of COVID-19 hospitalization and of ICU treatment.

#### Mandatory notification and reporting system

The mandatory notification and reporting system in Germany covers PCR-confirmed SARS-CoV-2 infections. These notifications fulfill the case definition of a COVID-19 case, regardless of information on symptoms and hospitalization status. The information on each COVID-19 case is sent from the laboratory to the local health authority (LHA) in charge. The LHA has to investigate all further epidemiological information. From the LHA, all notified COVID-19 case information is transferred via the health authorities of the federal states to the Public Health Institute. For our analysis, we used COVID-19 cases with known hospitalization as well as COVID-19 cases with known hospitalization and ICU admission (7, 14). For stratified analyses, we used age information from the notification system. Moreover, we used the information on known hospitalization status of all notified COVID-19 cases. We included all COVID-19 cases with known hospitalization and hospitalization dates from week 10/2020 to week 20/2022 or, if no hospitalization date was reported, with reporting dates from week 10/2020 to week 20/2022. Because of the broader case definition in the mandatory notification system, hospitalized COVID-19 cases could be either hospitalized due to a SARS-CoV-2 infection or for other reasons but with a confirmed SARS-CoV-2-infection.

#### Federal Statistical Office

From the full population data by the Federal Statistical office, we used the annual population and the number of discharged patients of the years 2015 to 2019, stratified by age, sex and federal state (15). These data were used for the estimation of the catchment population of the ICOSARI sentinel. For the validation of the SARI incidence estimation based on SARI cases from the 71 sentinel hospitals, we used full population data on cases with SARI diagnosis (ICD-10 codes J09 to J22) by the Federal Statistical Office as well as annual population data of the years 2015 to 2019. These data are available only with a time lag of approximatly one year and not suitable for a timely analysis. For incidence calculations, age-stratified annual population data of the years 2015 to 2021 were used. For the year 2022, we assumed the values of 2021.

Data from the ICOSARI sentinel and from the mandatory system were extracted on 28/July/2022, data from the Federal Statistical Office were extracted on 04/July/2022.

### Necessary preliminary analyses for the main study

#### Estimation of yearly catchment population of the ICOSARI sentinel hospitals

Using the ***denominator dataset*** on all-cause admissions, we estimated the yearly catchment population of the sentinel hospitals. We applied the proportional flow method as proposed by Norris and Bailey (16-18). We defined strata_*ijk*_ using age groups (*i* = 1,…,19; below 1 year, 1 to 4 years, 5 to 9 years, …, 85 to 89 years, 90 years and above), sex (*j*= 1,2; female, male) and regions (*k*= 1,…,4; North/West, Central/West, East, South; each region comprising more than one federal state). Within each stratum, we calculated the proportion of yearly sentinel patient admissions among national yearly patient admissions from full population data by the Federal Statistical Office (15). We derived the yearly stratum catchment population *CP*_*ijk*_ by multiplying the resulting proportion by the national population from the respective year.

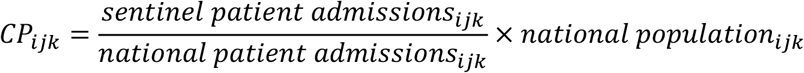

To derive the yearly catchment population *CP*_*i*_ of age groups, we summed up the yearly stratum catchment populations *CP*_*ijk*_ over regions and sex.

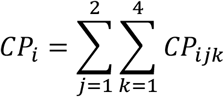

To derive the yearly total catchment population *TCP* of the sentinel, we summed up the catchment populations *CP*_*i*_ of all age groups. Age groups comprising larger age ranges were calculated accordingly by summing up the catchment populations 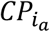 of age groups *i*_*a*_ that were within age group *a*.

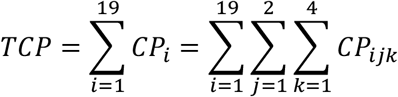

We estimated the yearly total catchment population *TCP* of the ICOSARI sentinel for the years 2015 to 2019 as described above. During the COVID-19 pandemic, the number of patient admissions (within the sentinel as well as country-wide) was affected by pandemic control and management measures and could not be used accordingly. Thus, we used the median stratum catchment population 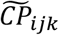 of the yearly *CP*_*ijk*_ from 2015 to 2019 to obtain the yearly catchment population 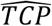 for the years 2020 to 2022.

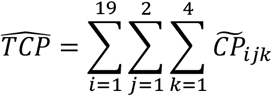

#### Estimation of weekly hospitalization incidence for SARI

We estimated weekly incidences of SARI by using the weekly number of newly admitted patients in the sentinel (numerator) and the yearly total catchment population of the sentinel. We validated our method by comparing weekly SARI incidence estimations from the ICOSARI sentinel to the published SARI incidence derived from full population data by the Federal Statistical Office using primary diagnoses of the years 2015 to 2019 (15). We described the fit graphically and by describing the weekly difference of the estimated and the actual SARI incidence. We calculated median and interquartile range (IQR) of the weekly difference and obtained the median percentage of under- or overestimation. Additionally, we quantified the fit by calculating the median difference of estimated and actual incidence overall and calculated the median percentage of under- or overestimation.

### Main analysis: Estimation of weekly COVID-SARI hospitalization and ICU incidence and comparison with weekly COVID-19 hospitalization and ICU data from the mandatory notification system

We estimated the incidence of COVID-SARI and of COVID-SARI with treatment in intermediate or intensive care units (***COVID-SARI ICU***) as described above, using the weekly number of COVID-SARI and of COVID-SARI ICU cases with admission dates from week 10/2020 to week 20/2022 from the ICOSARI sentinel and the corresponding catchment population (19). We used these estimations to evaluate hospitalization and intensive care burden of COVID-19 from notification data during five COVID-19 waves. We assessed and described potential underreporting of hospitalization using the proportion of notified cases with unknown hospitalization status at the peak of each wave (14). We compared estimated hospitalization and intensive care incidences in 6 different age groups that have been established within the ICOSARI hospital sentinel (0 to 4 years, 5 to 14 years, 15 to 34 years, 35 to 59 years, 60 to 79 years, 80 years and above). We described the fit graphically and stated the absolute difference (Δ_*a*_= *incidence*_*ICOSARIsentinel*_−*incidence*_*notificationdata*_) as well as the relative change (Δ_*r*_ = Δ_*a*_/*incidence*_*ICOSARIsentinel*_) of the incidence estimations at the peak of each wave. The five waves were classified as denoted in table 1 (20, 21).

**Table 1:** COVID-19 waves in Germany from week 10, 2020 to week 20, 2022. *wave still ongoing in week 20/2022

| Period | Start of period (week) | End of period (week) |
| --- | --- | --- |
| **First Wave** (wild type) | 10/2020 | 20/2020 |
| Period of low circulation | 21/2020 | 39/2020 |
| **Second Wave** (wild type) | 40/2020 | 8/2021 |
| **Third Wave** (VOC Alpha) | 9/2021 | 23/2021 |
| Period of low circulation | 24/2021 | 30/2021 |
| **Fourth Wave** (VOC Delta) | 31/2021 | 51/2021 |
| **Fifth Wave** (VOC Omicron) | 52/2021 | * |

## Results

### Estimation of catchment population of the ICOSARI sentinel hospitals

In the years 2015 to 2019, the yearly number of patient admissions within the ICOSARI hospital sentinel was 1.1 million. The number of admissions was markedly lower in the years 2020 and 2021. We thus estimated a total sentinel catchment population *TCP* of 4.4 to 4.5 million for the years 2015 to 2019 and obtained a total catchment population 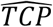of 4.4 million, which we used as denominator for the years 2020 to 2022 (see table 2).

**Table 2:** Number of yearly patient admission in the ICOSARI sentinel and estimated catchment population.

| Year | Number of patient admissions in sentinel | Estimated catchment population in sentinel |
| --- | --- | --- |
| 2015 | 1,106,904 | 4,406,215 |
| 2016 | 1,116,127 | 4,403,521 |
| 2017 | 1,108,775 | 4,405,180 |
| 2018 | 1,108,153 | 4,448,800 |
| 2019 | 1,110,341 | 4,463,590 |
| 2020 | 960,619 | 4,423,022 |
| 2021 | 925,195 | 4,423,022 |
| 2022 | 366,770 | 4,423,022 |

Figure 1 shows the weekly estimated SARI incidence per 100,000 population in the ICOSARI sentinel compared to the incidence retrospectively derived from the full population data by the Federal Statistical Office. Using the sentinel data, we overestimated the weekly SARI incidence by less than 1 SARI case per 100,000 population (Median [IQR]: 0.97 [0.45;1.69]), with larger differences in the peak weeks of the seasonal influenza epidemic (Figure 1). That corresponds to an overestimation of 10% in median. Overall, the overlap of sentinel data and the full population data was very good regarding the timing and the level of trends and peaks.

**Figure 1:**
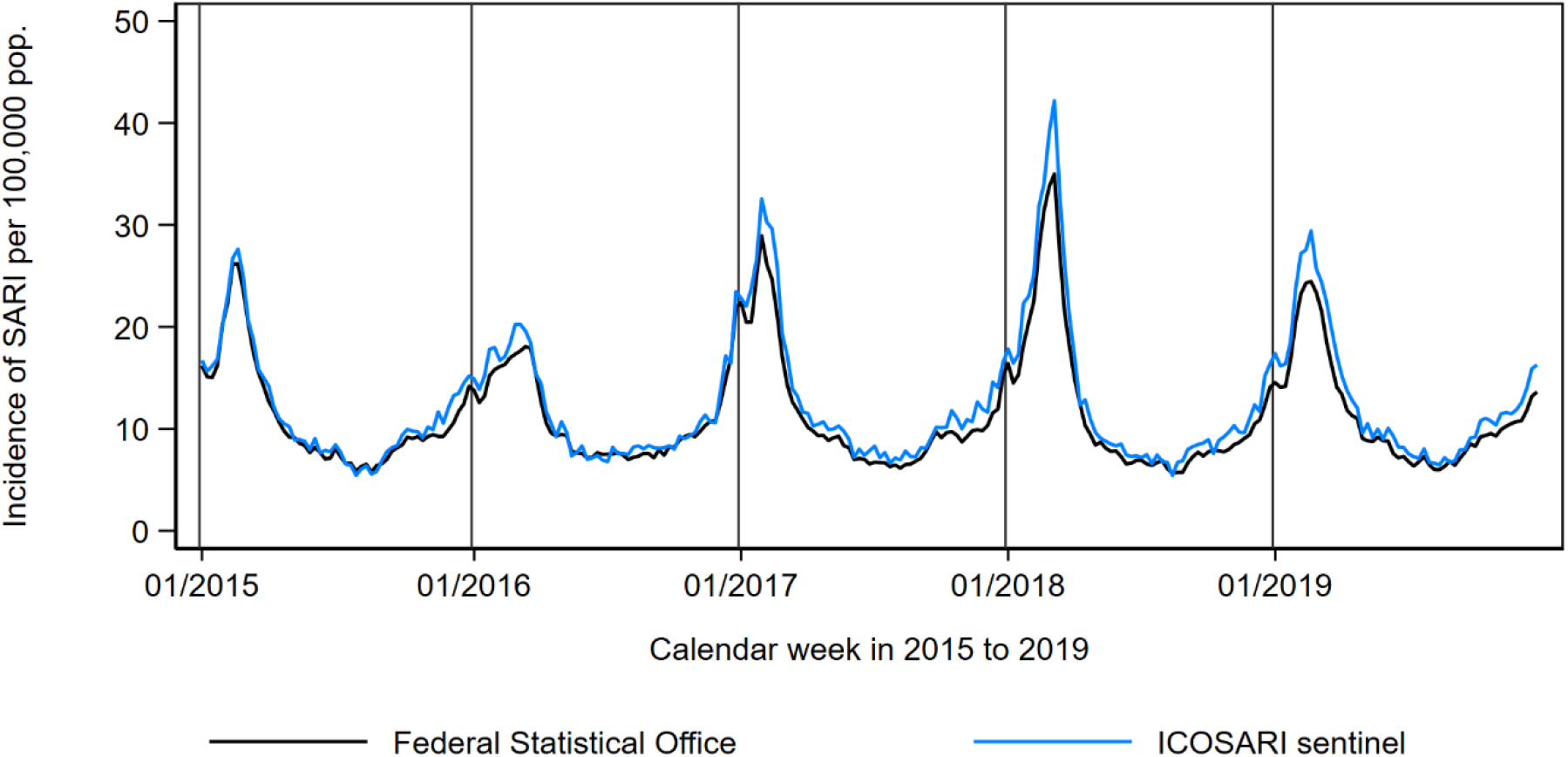
Estimated incidence of SARI cases (ICD-10 codes J09 to J22), using ICOSARI sentinel data and full population data from the Federal Statistical Office by admission week from 1/2015 to 52/2019.

### Comparison of COVID-SARI hospitalization and ICU incidence with “all cause” COVID-19-hospitalization and ICU incidence from mandatory notifications

From week 10/2020 to week 20/2022, there were 26,798 COVID-SARI cases and 7.335 COVID-SARI ICU cases in the ICOSARI sentinel. Notification data comprised 556,400 hospitalized COVID-19 cases and 51,857 cases receiving intensive care.

During the first wave (**peak 1** - week 13/2020: 6.6 vs 6.2 per 100,000 population, Δ_*a*_ = 0.4 per 100,000), the estimated hospitalization incidences matched with only 6% relative change Δ_*r*_ of COVID-SARI incidence compared to COVID-19 hospitalization incidence from notification data (Figure 2). However, during the second, third and fourth wave (**peak 2** - week 52/2020: 21.9 vs 13.4 per 100,000 population, Δ_*a*_ = 8.5 per 100,000; **peak 3** - week 14/2021: 13.7 vs. 9.2 per 100,000 population, Δ_*a*_ = 4.5 per 100,000; **peak 4** - week 46/2021: 15.9 vs. 12.7 per 100,000 population, Δ_*a*_ = 3.2 per 100,000), the COVID-SARI incidence assumed much higher values than the COVID-19 hospitalization incidence from notification data with 39%, 33% and 20% relative change Δ_*r*_ at the peak weeks. Throughout the fifth wave, the COVID-SARI incidence was lower than the COVID-19 hospitalization incidence from notification data (**peak 5** - week 11/2022: 10.6 vs. 14.7 per 100,000 population, Δ_*a*_ = -4.1 per 100,000) with Δ_*r*_ = -39% at the peak.

**Figure 2:**
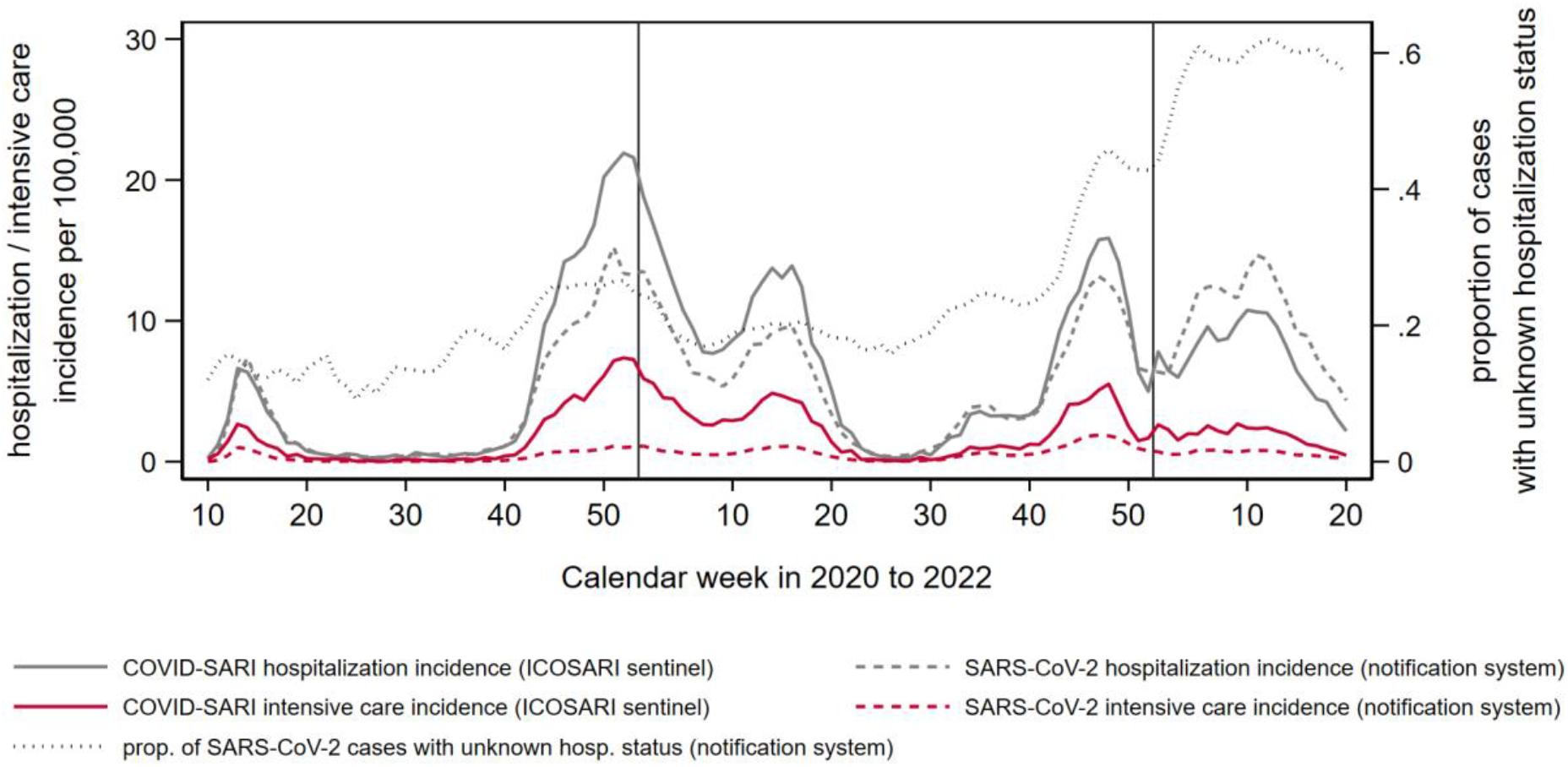
Estimated hospitalization incidence and intensive care incidence: COVID-19 cases with diagnosis code for SARI (ICD-10 codes J09-J22) from the hospital sentinel ICOSARI (solid line), for hospitalized COVID-19 cases from notification data (dashed line); proportion of COVID-19 cases with unknown hospitalization status from notification data (dotted line)

The COVID-SARI incidence from sentinel data indicated, that the second wave had the highest peak burden of severe COVID-19 cases, followed by the fourth, the third, the fifth and the first wave. However, the COVID-19 hospitalization incidence from notification data assumed similar peak values in the second and the fifth wave, followed by the fourth, the third and the first wave. Both estimations corresponded in having the lowest peak values during the first wave.

Within the notification data, the proportion of COVID-19 cases with unknown hospitalization status was 15% at **peak 1**. In the later waves however, the proportion was much higher with 27% and 20% at **peak 2** and **peak 3**, respectively. It increased further to 46% in **peak 4** and 61% in **peak 5**.

The estimated incidence of COVID-SARI ICU from sentinel data was more than twice as high as the estimated incidence of COVID-19 ICU admissions derived from notification data (Figure 2). The COVID-SARI ICU incidence peaked at values 2.7, 7.4, 4.9, 5.5 and 2.4 per 100,000 in the five waves with similar peak values during the first and the fifth wave, whereas the highest value overall at **peak 2** was three times higher. In contrast, the COVID-19 ICU incidence from notification data remained well below 2.0 per 100,000 for all five peaks (1.0, 1.0, 1.1, 1.8 and 0.8 per 100,000 population), with very similar peak values in four of the five waves. The relative changes Δ_*r*_ ranged between 63% (at **peak 1**) and 86% (at **peak 2**). This considerable difference in ICU incidence estimations is also reflected by the proportion of ICU admissions among COVID-19 hospitalizations, which was 40%, 34%, 35%, 35% and 22% for the COVID-SARI patients in the sentinel, but only 17%, 7.6%, 12%, 14 and 5,5% in the notification data at the **peaks 1** to **5**. The proportion of ICU admissions was highest at **peak 1** and lowest at **peak 5** for COVID-SARI as well as for COVID-19 hospitalization from notification data. Looking at the data from the ICOSARI sentinel, the proportion of ICU admissions among COVID-SARI patients was very similar at the peaks **2, 3** and **4** (34% to 35%). However, looking at the notification data, the proportion of ICU among COVID-19 hospitalizations was much lower at **peak 2** (7.6%) compared to the **peaks 3** and **4** (12% and 14%).

The estimation of COVID-19 hospitalization incidence by age groups is shown in Figure 3. For the age groups below 35 years, the level of COVID-SARI hospitalization incidence from the ICOSARI sentinel was consistently lower than the level of COVID-19 hospitalizations from notification data. In the age groups 0 to 4 years and 5 to 14 years, the first three waves were not reflected in the incidence from the ICOSARI sentinel with values well below 1 COVID-SARI hospitalization per 100,000 population. However, the fourth and fifth wave showed slightly larger values in children, but still remained low with values well below 3 COVID-SARI hospitalization per 100,000 population.

**Figure 3:**
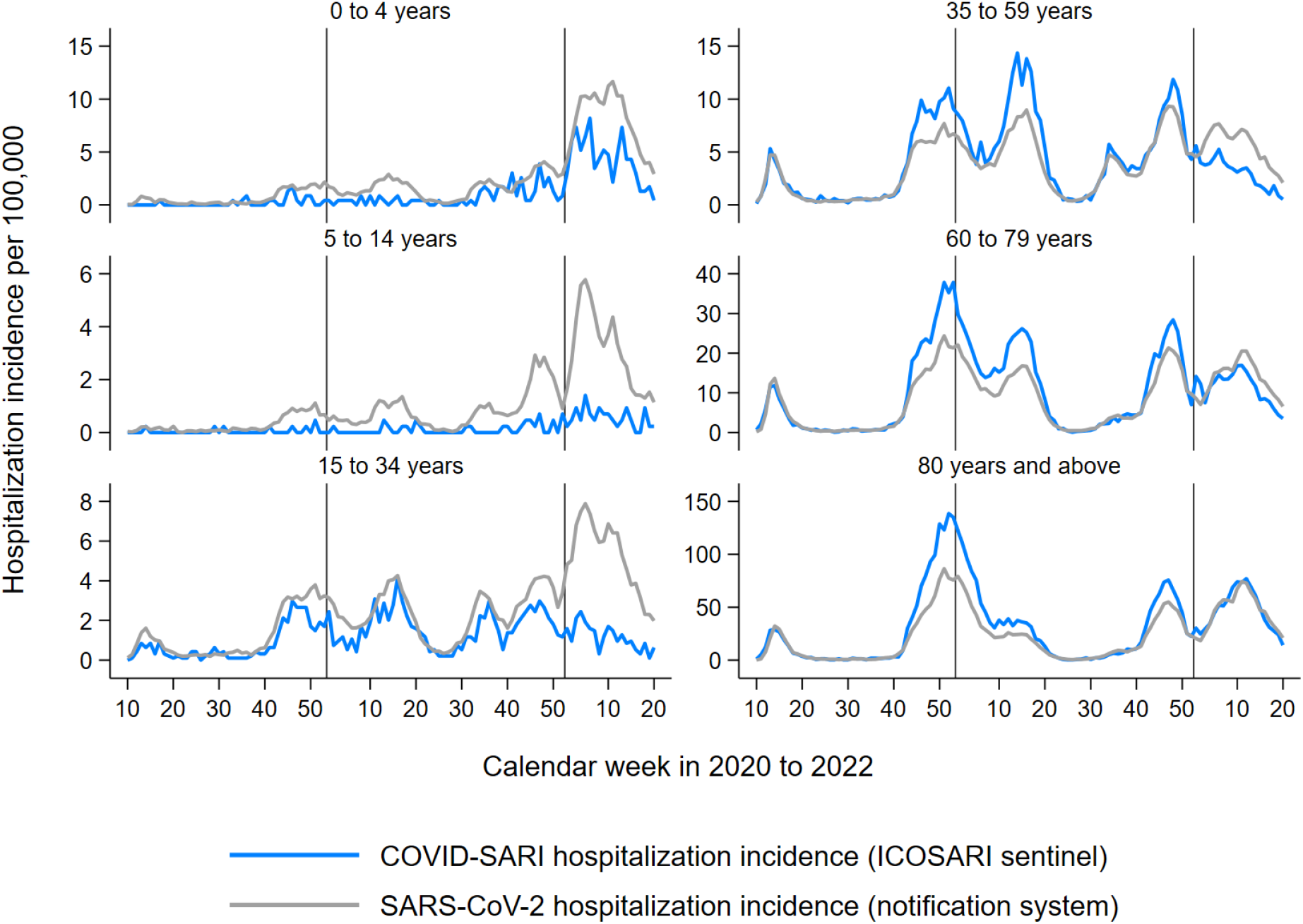
Estimated hospitalization incidence by age groups: COVID-19 cases with diagnosis code for SARI (ICD-10 codes J09-J22) from the hospital sentinel ICOSARI (blue) and for hospitalized COVID-19 cases from notification data (gray)

Using notifications data, the COVID-19 hospitalization incidence estimation for children below 15 years reflected all five waves. However, it showed a similar pattern as the COVID-SARI incidence from the ICOSARI sentinel, with incidences below 3 per 100,000 population in the first three waves and higher incidences in the fourth and fifth wave (up to 12 per 100,000 in the fifth wave for children below 5 years). In the age groups 15 years an older, all waves were well reflected in the COVID-19 hospitalization incidences both from the ICOSARI sentinel data and the notification data. In the age groups 35 years or older, the match in incidence estimations from the two data sources was good in the first wave with of 11%, -6% and 25% relative change Δ_*r*_ at **peak 1**, respectively. The match was less in the second and the third wave with Δ_*r*_ of around 40% at the peaks, but improved again in the fourth wave (Δ_*r*_ between 22% and 28% at **peak 4**). In the fifth wave, the estimated COVID-SARI incidence from the ICOSARI sentinel was below the COVID-19 hospitalization incidence for the age groups 35 to 59 years (Δ_*r*_= -112% at **peak 5**) and 60 to 79 years (Δ_*r*_= -21% at **peak 5**). In the age group 80 years and above, the estimations from both data sources were very similar with a relative change Δ_*r*_ of -1% at **peak 5**. Both data sources revealed the highest hospitalization incidences in the age group 80 years and above for all of the five waves.

The estimation of COVID-19 ICU incidence by age groups is shown in Figure 4. For the age groups below 15 years, the incidence level was consistently below 0.5 COVID-19 ICU patients per 100.000 from both data sources and the waves were not reflected in the ICU incidences. In the age group 15 to 34 years, only the second, third and fourth wave were reflected in the COVID-19 hospitalization incidences both from the ICOSARI sentinel data and the notification data. For all age groups 35 years and older, the ICU incidence of COVID-SARI cases from the ICOSARI sentinel was markedly higher than the ICU incidence of COVID-19 cases from notification data with relative changes Δ_*r*_ of at least 50% at the peaks. These differences were equally large for all five waves. Both data sources showed the highest the ICU incidences in the age group 80 years for most waves, but found higher ICU incidences in the age group 60 to 79 years during the third COVID-19 wave.

**Figure 4:**
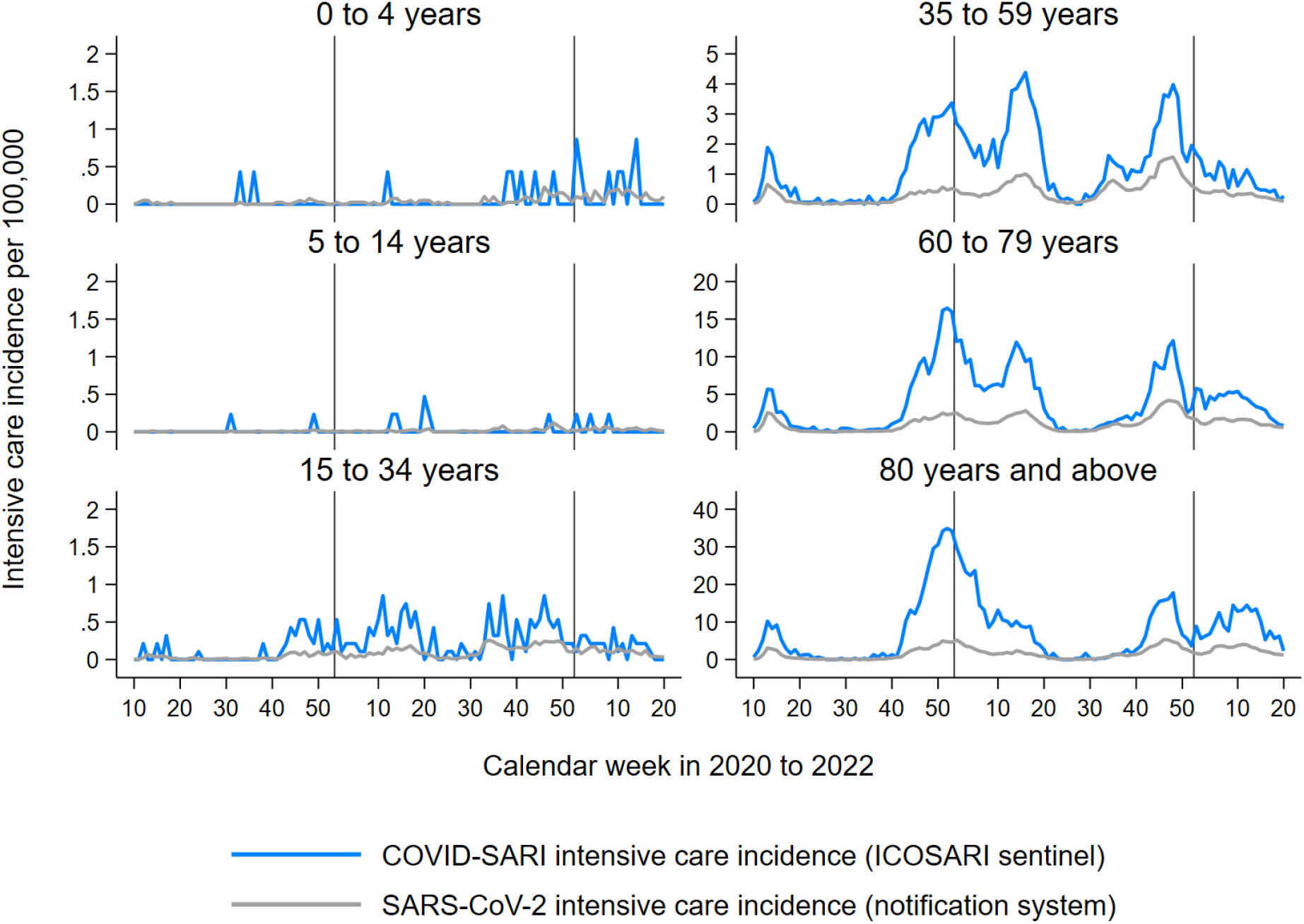
Estimated intensive care incidence by age groups: COVID-19 cases diagnosis code for SARI (ICD-10 codes J09-J22) from the hospital sentinel ICOSARI (blue) and for hospitalized COVID-19 cases from notification data (gray)

## Discussion

We estimated weekly hospitalization and intensive care incidences using syndromic hospital data of SARI cases. The burden of hospitalized COVID-19 cases differed when using the data on SARI cases compared to the notification data. Regarding the COVID-SARI hospitalization and ICU incidence from ICOSARI sentinel, the second wave with circulating SARS-CoV-2 wild type had a distinctly higher burden overall at the peak week with at least 38% more cases than the other waves at the peak week. This can largely be attributed to very high COVID-SARI incidences in the age groups 60 years and older during the second wave. However, no distinctly high hospitalization peak burden could be derived from notification data in the second wave compared to the other waves and the COVID-19 incidences from notification data in the age groups 60 years and older were similar during the second, fourth and fifth wave.

With the emergence of the Omicron VOC in the fifth wave, the importance to differentiate between hospitalizations with COVID-19, as recorded in the notification data, and hospitalizations due to COVID-19, as captured by COVID-SARI cases from ICOSARI sentinel data, becomes even more pronounced. This is clearly illustrated in the relative change Δ_*r*_, which indicated underestimation of COVID-19 hospitalization at the peak of the second, third and fourth wave with values ranging from 20% to 39%, but showed considerable overestimation at the peak of the fifth wave with Δ_*r*_ of -39%. In capturing all laboratory confirmed SARS-COV-2 infections with known hospitalization regardless of symptoms, the notification system uses a broader case definition for COVID-19 hospitalizations (“all-cause” hospitalizations with COVID-19) than the ICOSARI sentinel, which captures COVID-SARI (hospitalizations due to severe COVID-19). Thus, we attribute the overestimation during the fifth wave to a lower proportion of hospitalizations due to severe COVID-19 (COVID-SARI) among all COVID-19 hospitalizations and consequently a higher proportion of hospitalizations for other reasons than the laboratory confirmed SARS-CoV-2 infection (i. e. “with COVID-19”) during the Omicron wave compared to previous waves. This is in line with other findings internationally, where a high overall incidence was observed, but a significantly lower disease severity regarding hospitalization, ICU treatment and fatality was found (22-26). The markedly lower proportion of ICU patients among COVID-SARI hospitalizations at the peak of the fifth wave (22%) compared to the previous peaks (34 to 40%), as noted in the ICOSARI sentinel data, confirms these findings.

Other studies have pointed out, that although there was a reduction in disease severity during the Omicron wave, it is less pronounced in the elderly (22, 24-26). Our analysis supports this, as COVID-SARI hospitalization incidence from sentinel data and COVID-19 hospitalization incidence from notification data overlapped in the age group 80 years and older during the fifth wave (relative change Δ_*r*_= -1%). In all other age groups, the “all-cause” hospitalization incidence with COVID-19 (notification data) was higher than the hospitalization incidence due to severe COVID-19 (ICOSARI sentinel data).

Prior to our analysis, we expected consistently higher COVID-19 hospitalization incidences from notification data with the broader case definition compared to COVID-SARI hospitalization incidences, where only cases with severe respiratory symptoms are captured. Considering the increasingly high proportion of unknown hospitalization status during the waves, we assume that underreporting of COVID-19 hospitalizations especially during peak times in the later waves (after the first wave) and of intensive care treatments in general was present in the notification data. During the Omicron wave, we interpret the high hospitalization incidences with COVID-19 from notification data as reflecting the very high SARS-CoV-2 incidence in the general population rather than the true burden of severe illness due to COVID-19. Thus, although there might still have been underestimation of hospitalizations with COVID-19 in the notification data, we assume an overestimation of severe COVID-19 burden during the Omicron wave.

Our analysis shows that the hospital sentinel provides robust data on COVID-SARI in times when the pressure on the health system is very high, as was observed especially during the second COVID-19 wave at the end of 2020. Several other countries in Europe and also on other continents had waves with very high case numbers at the end of 2020, followed by a weaker wave in early 2021. Remarkably, hospitalization and intensive care incidences in the later wave (early 2021) were lower in the older age groups compared to the wave at the end of 2020, whereas younger age groups tended to have even higher incidences of severe cases in the early 2021 wave than in the previous wave (27-30). The reduced burden in the oldest age group is consistent with the start of the immunization program in Germany at the end of 2020, which mostly prioritized older age groups and effectively prevented severe cases in the most vulnerable population (31).

The rate of ICU among COVID-SARI patients in the ICOSARI sentinel is representative for German hospitalized COVID-19 patients and aligns with other sources for the first wave in Germany as well as in other countries (32-35). The gap in the intensive care incidence estimated from notification data and from sentinel data can largely be attributed to underreporting in the notification data. To some extent however, the definition of COVID-SARI may contribute to a higher ICU rate, as it includes only cases with severe respiratory infections and excludes COVID-19 cases without respiratory symptoms that were detected via admission screening in hospitals. However, both data sources consistently showed plausible trends regarding the differences in burden between age groups.

The strength of the hospital sentinel system is the daily electronic submission of quality-assured case-based data from hospital to national health institute. The utilization of ICD-10 data is a digital standardized method without additional workload for the hospitals. It is both timely and enables international comparability. Thus, the system provides validated, complete and conclusive data also in times of high pressure on the health system, where local health authorities may be challenged with follow-up of COVID-19 cases, which is time and resource intensive. Moreover, the syndromic surveillance system provides several years of SARI time series, allowing comparisons of COVID-SARI burden with the SARI burden during influenza waves (13, 33, 36).

However, our findings are subject to some limitations. The estimation of regional catchment population with the given data is difficult, therefore larger regions based on federal states were used (15). Yet, regional estimations are a strength of notification data.

For the years 2020 and 2021, we saw a notable decrease of patient admissions in the hospital sentinel. This effect was observed worldwide and was probably due to a combination of aspects such as the cancelling of elective operations and an increased hesitancy to use the health service in general and especially hospitals (37-44). As we had stable catchment population estimations in the preceding years, using a fixed catchment population derived from the median of previous years was justified.

The number of COVID-19-cases detected in hospitals is dependent on testing, and is likely underestimated. However, routine screening upon admission in sentinel hospitals was in place since July 2020. Furthermore, cases with laboratory confirmed SARS-CoV-2 infection hospitalized due to symptoms other than severe acute respiratory infections are not reported within the SARI sentinel. Thus, the focus on COVID-19 cases with SARI is more robust and less biased by the testing strategy as seen in notification data, as pneumonia was known to be the main syndrome of severe COVID-19 since the first case reports from China (1). We also note that weekly estimation of age-specific hospitalization and ICU incidences can lack accuracy due to low case numbers, especially in the younger age groups.

## Conclusion

A valid link between the total number of notified COVID-19 cases, hospitalized patients and ICU patients is essential for the assessment of the seriousness of COVID-19, particularly considering the repeated emerging of new concerning variants (4, 36). Using data from the syndromic surveillance system ICOSARI allows robust and specific estimations of hospitalization incidence due to severe COVID-19, adds important information on intensive care burden in COVID-19 patients and thus enables to evaluate the different severity of the five COVID-19 waves in Germany. Especially in times of high caseloads, the investigation of cases by local health authorities may lead to incomplete data on the severity of cases. In contrast, at that period the SARI sentinel provided robust estimations on the proportion of intensive care and on intensive care incidence. Moreover, data from the syndromic ICD-10 based SARI sentinel is well suited to assess the true burden of severe COVID-19 disease, where all-cause hospitalizations with SARS-CoV-2 laboratory confirmation might overestimate the burden. Thus, SARI surveillance is an important instrument to assess disease seriousness and to provide a timely and robust evidence base for decisions on preventive measures, additionally to the monitoring of transmission and of capacity utilization (2, 45-47). Importantly, the use of an existing system has many advantages such as known stability and potential biases, baseline data from previous years as well as known catchment population which is essential for the estimation of burden.

The method for the estimation of hospitalization and intensive care incidence from syndromic sentinel data can be applied not only in the COVID-19 pandemic (48). In particular, continuous surveillance of severe cases is essential to accompany the transition from a pandemic to an endemic disease. Moreover, syndromic ICD-10 based SARI surveillance is a sustainable tool for seasonal epidemics such as seasonal influenza and RSV disease and also an essential component for pandemic preparedness of respiratory viral diseases, as it can be further adapted to other respiratory case definitions if needed (48, 49). To improve regional representativeness, stepwise expansion of the system is planned.

## Data Availability

All data produced in the present study are available upon reasonable request to the authors.

## Notes

### Competing Interest Statement

The authors have declared no competing interest.

### Funding Statement

The work of Kristin Tolksdorf was funded as a research project by the German ministry of health.

### Author Declarations

Ethics committee of Aerztekammer Berlin KdoeR waived ethical approval for this work.

### Summary of Updates

analyzed time period extended to include circulation of VOC Omicron; methods section restructured and extended; results section revised; discussion section reorganized

